# Accelerated high-capacity 6 mA transcranial direct current stimulation for major depression

**DOI:** 10.64898/2026.10.05.26364655

**Authors:** Clayton Olash, Elisabeth Collins, Danyal Smani, Stewart S. Cox, Marom Bikson, Kyle Donnery, Mohamad FallahRad, Mojtaba Belali Koochesfahani, Jean-Philippe Miron, Crawford Latham, Dylan Freels, Joe Chasen, Mark S. George

**Affiliations:** Brain Stimulation Laboratory, Neuromodulation and Behavioral Medicine Division, Department of Psychiatry and Behavioral Sciences, Medical University of South Carolina, Charleston, SC, USA; Department of Biomedical Engineering, The City College of New York, City University of New York, New York, NY, USA; Interventional Psychiatry Program, UC San Diego Health, San Diego, CA, USA; College of Medicine, Medical University of South Carolina, Charleston, SC, USA; Ralph H. Johnson VA Medical Center, Charleston, SC, USA

## Abstract

**Objective:** Home-based 2 mA transcranial direct current stimulation (tDCS) is FDA-approved for depression, but amplitude has been limited largely by skin tolerance. This study evaluated the safety, tolerability, and feasibility of accelerated high-capacity tDCS (HC-tDCS) at 6 mA.

**Methods:** In this open-label pilot study, 20 patients with major depressive disorder received 20-minute bifrontal 6 mA sessions twice daily for five days using a high-capacity stimulator and hydrogel electrodes. A Safety Review Committee reviewed data from the first three participants before the remaining 17 were enrolled.

**Results:** Retention and adherence were 100% (203 of 203 sessions); 17 of 20 received 6 mA throughout, with amplitude reduced for intolerance in one and limited by an electrode fault in two. There were no serious adverse events. The most common symptoms were fatigue (49% of sessions), burning (46%) and tingling (43%) at the electrode, and difficulty concentrating (40%). Electrode-site erythema occurred in all participants, with no severe reactions. At 1 week, 7 of 20 reported a residual skin change, none painful; at 1 month, spots of residual pigmentation were observed at the electrode site in 2 participants, both Black. In five days, mean PHQ-9 scores decreased from 15.8 to 7.2 (Hedges g=1.62); 50% of participants responded, 45% remitted, and improvement was maintained at 1 month. Anhedonia, mindfulness, and self-reported cognitive function also improved (g=0.76 to 1.23), with gains maintained at 1 month.

**Conclusions:** Accelerated 6 mA HC-tDCS was feasible, well tolerated, and warrants a sham-controlled trial.

## Introduction

Depressive disorders affect an estimated 332 million people worldwide and are strongly associated with suicide ^1,2^. Only about one-third of patients achieve remission with initial antidepressant pharmacotherapy, and those requiring successive agents accumulate a substantial side-effect burden, highlighting the need for more effective, rapid-acting, and accessible therapies ^3^. Brain stimulation treatments, such as electroconvulsive therapy (ECT) and transcranial magnetic stimulation (TMS), achieve response or remission in approximately half of patients with treatment-resistant depression (TRD) ^4–7^. But these modalities are expensive, resource-intensive, have associated risks, and require repeated visits to a specialized center, which limits their accessibility. In a national Veterans Affairs cohort, fewer than 1% of patients with depression or PTSD received interventional psychiatric treatment ^8^.

Transcranial direct current stimulation (tDCS) delivers a low-intensity constant current, conventionally 1-2 mA between two scalp electrodes. tDCS is relatively inexpensive and portable, even self-administered at home ^9^. In December 2025, the US Food and Drug Administration (FDA) approved home-delivered 2 mA tDCS for depression, but the approved course takes 10 weeks and is limited to patients “not considered treatment refractory to medication”^10^. Patients with TRD respond poorly and inconsistently to conventionally dosed tDCS ^11^. There therefore remains a need for a scalable intervention that acts within days and remains effective even in TRD.

High-Capacity tDCS (HC-tDCS), which combines engineered single-use hydrogel electrodes with adaptive current controls, allows for high intensity (e.g., 6 mA) per session and/or sessions per day, by reducing adverse skin responses ^12^. In healthy subjects, HC-tDCS at 6 mA was as well tolerated as 2 mA sponge tDCS, while producing >4x brain electric fields ^12^.

Several lines of evidence suggest that increasing tDCS intensity may enhance or accelerate clinical effects. The conventional 1-2 mA dose was inherited from acute motor-cortex TMS physiology ^13,14^ rather than derived by therapeutic dose-finding, and has persisted for over 20 years, largely because of skin tolerability concerns with conventional sponge electrodes ^15^. Current delivered to the brain (electric field) increases linearly with current intensity ^16,17^, and cellular markers of response, including polarization, firing rate, oscillation power, and plasticity, increase monotonically with electric fields ^18,19^. Because variations in individual anatomy result in different electric fields for a fixed applied current, several retrospective analyses correlated individual electric fields with clinical or cognitive response ^20–23^.

Both stimulation intensity and cumulative exposure influence tDCS antidepressant effects ^9,11,24–26^. In addition to total dose, accelerated sessions – dosing multiple times within a day – may be important ^24,27^. This parallels developments in TMS, where accelerated protocols, such as Stanford Accelerated Intelligent Neuromodulation Therapy (SAINT), compress weeks of daily treatment into multiple daily sessions over just five days ^28,29^. Thus, with tDCS, as with other brain stimulation modalities, increasing intensity and accelerated protocols may prove more effective, faster, or both.

In this study, we evaluated accelerated HC-tDCS, 6 mA delivered twice daily over five days in 20 patients with major depressive disorder, three-quarters of whom reported treatment resistance. The study was designed to assess safety, tolerability, and feasibility of accelerated HC-tDCS while exploring its potential for rapid antidepressant and neurocognitive effects.

## Methods

### Study overview

This was a single-arm, open-label, outpatient pilot study conducted at the Brain Stimulation Laboratory of the Medical University of South Carolina (MUSC), Charleston, South Carolina. The protocol was approved by the MUSC Institutional Review Board (Pro00146983) and registered at ClinicalTrials.gov (NCT07226011). Written informed consent was obtained from all participants, with electronic consent available for participants unable to attend the laboratory beforehand. The study was conducted under a non-significant-risk device determination. Reporting follows the CONSORT 2010 extension for pilot and feasibility trials, adapted for a single-arm design.

The design incorporated a two-stage safety architecture. The first three participants completed a within-day escalation to 6 mA on Day 1 before continuing on the standard schedule (Fig. 1B). An independent Safety Review Committee reviewed their adverse-event, skin and tolerability data and determined whether continuation at 6 mA was acceptable before the remaining participants were enrolled at 6 mA from Day 1. No quantitative criteria for progression to a definitive trial were prespecified.

**Figure 1.**
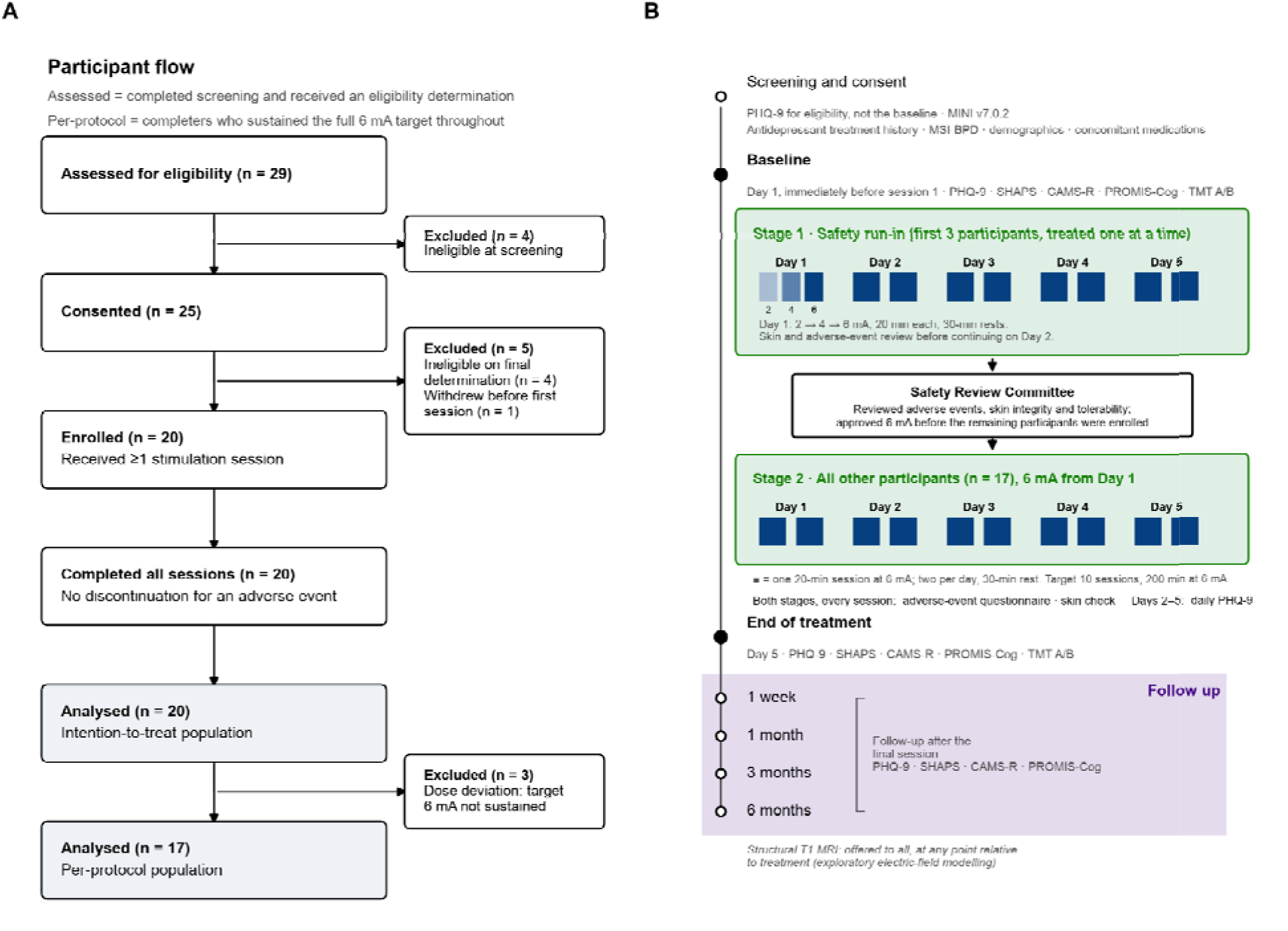
Study flow and schedule. (A) Participant flow from eligibility assessment to the analysis population; individuals assessed for eligibility completed the screening instrument and received an eligibility determination, and those referred but never screened are not shown. (B) Assessment and stimulation schedule. The first three participants completed a safety run-in with a within-day escalation to 6 mA on day 1 (stage 1); a Safety Review Committee reviewed their adverse-event, skin-integrity and tolerability data before the remaining 17 participants were enrolled at 6 mA from day 1 (stage 2). Filled squares denote 20-minute sessions, two per day.

### Participants

Adults aged 18–70 years were eligible if they had a current diagnosis of major depressive disorder established by the Mini-International Neuropsychiatric Interview version 7 (MINI v7.0.2), a baseline PHQ-9 score greater than 9, capacity to provide informed consent, and spoke fluent English. Key exclusion criteria were bipolar or psychotic disorder, current suicidality requiring a higher level of care, neuromodulation therapy within the preceding six months, seizure risk factors, and any skin abnormality at the electrode sites; the full criteria are given in Supplementary Methods 1.

Participants were recruited through referrals from the MUSC brain stimulation and interventional psychiatry services and campus advertising; the planned primary-care pathway yielded no referrals. Age, sex assigned at birth, gender identity, ethnicity and race were self-reported on a standardized demographic form using the U.S. Office of Management and Budget race and ethnicity categories, with more than one race permitted.

### Stimulation protocol

Stimulation was delivered with an investigational High-Capacity 1×1 stimulator (HC-1×1; Soterix Medical) with a maximum output of 6 mA, through single-use, self-adhesive high-capacity hydrogel electrodes (25 cm²; laboratory of M.B., City College of New York) replaced for every session. Electrodes were placed in a bifrontal montage, anode over the left and cathode over the right frontal region, each positioned as laterally as possible following the contour of the eyebrow (Supplementary Fig. 1). Device and electrode specifications are given in Supplementary Methods 1.

Each session delivered 6 mA for 20 minutes, twice daily with a 30-minute rest, on five consecutive weekdays, for a target of 10 sessions per participant; the three run-in participants received one additional Day 1 session, giving 203 sessions in total. All sessions were delivered in the laboratory under continuous staff monitoring, and any session in which 6 mA was not sustained was recorded by staff. No task or therapy was paired with stimulation.

#### Dose-reduction procedure

Participants unable to tolerate 6 mA had the amplitude reduced in stepwise decrements to an individualized tolerable level, carried forward for their remaining sessions. A reduction did not constitute discontinuation for the feasibility endpoint but defines the per-protocol population. In two participants, an electrode fault prevented the stimulator from reaching 6 mA; those sessions were delivered at the highest achievable amplitude.

### Outcomes

Primary outcomes were safety, tolerability and feasibility: (i) device-related serious adverse events, defined as events requiring hospitalization or emergency care during the stimulation period; (ii) the proportion of participants completing all scheduled sessions without discontinuation for adverse effects; and (iii) a tDCS Adverse Event Questionnaire adapted from Brunoni et al.^30^, administered after every session, on which each symptom is rated absent, mild, moderate, severe or extreme (scored 1 to 5). The Safety Review Committee determination after the run-in was an additional IRB-specified endpoint.

Before stimulation each day, staff inspected both electrode sites and recorded any finding and any action taken, such as repositioning the electrode; this check was introduced after the first run-in participant. After every session, staff graded erythema at the more affected site as none, mild, moderate or severe, and electrode sites were photographed before stimulation each day and after each session. At each follow-up visit, participants completed an electrode-site skin questionnaire and uploaded forehead photographs, which were reviewed by one unblinded investigator (C.O.), not a dermatologist (Supplementary Methods 1).

**Secondary outcomes** were change from baseline to Day 5 in depressive symptom severity (Patient Health Questionnaire-9, PHQ-9), anhedonia (Snaith–Hamilton Pleasure Scale, SHAPS), trait mindfulness (Cognitive and Affective Mindfulness Scale–Revised, CAMS-R), subjective cognitive function (PROMIS Cognitive Function 4a), and objective processing speed and executive function (Trail Making Test parts A and B); and change in depressive symptom severity from baseline to the 1 month follow-up. SHAPS and the Trail Making Test were prespecified secondary outcomes in the IRB-approved protocol but were not listed in the ClinicalTrials.gov registration. The self-report battery was repeated 1 week and 1 month after the final session. On the final day of treatment, a brief non-validated participant feedback questionnaire was given.

Clinician-rated HRSD-24 and MADRS assessments were added by protocol amendment partway through enrollment and administered at baseline and end of treatment; this was not a prespecified outcome. Treatment expectancy was rated from 0 to 10 before each session, and the rating before the first session was used. All participants were offered a structural MRI for electric-field modeling; modeled electric field in target regions is a registered secondary outcome and will be reported separately.

Baseline was the assessment on the first treatment day, immediately before the first session, and the end-of-treatment value was the final PHQ-9 of the treatment week. PHQ-9 recall windows and scoring conventions are given in Supplementary Methods 1. Study data were collected and managed using REDCap^31,32^.

### Analysis populations

Outcomes are reported in three nested populations: intention-to-treat (ITT), all participants who received at least one session; completers, whose sessions spanned five treatment days; and per-protocol, completers who sustained 6 mA throughout. ITT is the primary population for all outcomes, and safety is reported only on the ITT population.

### Statistical analysis

The study was designed to estimate safety, tolerability and feasibility and to generate effect-size estimates for a randomized trial; it was not powered to test efficacy, and the sample of 20 was chosen pragmatically. Without a sham arm, efficacy analyses describe within-participant change and cannot separate any effect of stimulation from expectancy, regression to the mean or the natural course of the episode. No statistical analysis plan was prespecified, and the response and remission thresholds were adopted after data collection.

Change from baseline to end of treatment was tested with two-sided paired t-tests, with Wilcoxon signed-rank tests as a sensitivity analysis, and summarized as Hedges’ g. Durability across baseline, end of treatment, one week and one month, and daily PHQ-9 within the treatment week, were analyzed with repeated-measures one-way ANOVA or, where data were missing, a mixed-effects model, with Greenhouse–Geisser correction as needed and Holm–Šídák post hoc comparisons against baseline. Response was defined as a reduction of at least 50% in PHQ-9 and remission as a score below 5; these and the safety proportions are reported with exact (Clopper–Pearson) binomial confidence intervals. Response by self-reported treatment resistance was compared post hoc with Fisher’s exact test. Adverse events and skin findings were tabulated descriptively. Methods for the expectancy and clinician-rated analyses are given in Supplementary Methods 1. All confidence intervals are 95%, and analyses were performed using GraphPad Prism (v11).

## Results

Between 15 December 2025 and 28 August 2026, 29 individuals were assessed for eligibility, 25 consented and 20 received at least one stimulation session, constituting the intention-to-treat population (Fig. 1). After the three run-in participants completed treatment, the Safety Review Committee determined that enrollment could proceed at 6 mA. All 20 completed the five-day course. Three did not sustain 6 mA throughout, one because of intolerance (amplitude reduced to 4.5 mA) and two because an electrode fault prevented the device from reaching 6 mA; all three were excluded from the per-protocol population (n=17). Mean age was 39.6 years (range 18 to 69), 11 of 20 were female at birth, and 15 (75%) reported two or more adequate prior antidepressant trials (Supplementary Table 1).

### Accelerated HC-tDCS was tolerated without severe skin reactions

No participant experienced a device-related serious adverse event (0 of 20; CI 0% to 17%), and all 20 completed every scheduled session without discontinuation for an adverse effect (100%; CI 83% to 100%). Electrode sites were graded at 200 of the 203 delivered sessions, and no severe skin reaction or skin breakdown occurred (Table 1); the highest grade recorded was mild in 12 participants and moderate in 8. Before stimulation on days 2 to 5, staff recorded an electrode-site finding, all but one mild and most often dryness or flaking, in 10 of 19 participants, and repositioned electrodes in 5. At one week, 7 of 20 participants reported a current skin change, most often discoloration (5) or flaking (4), and none reported tenderness or pain. At one month, 3 of 18 reported residual discoloration, judged on photographic review to be related to stimulation in 2 (Supplementary Results).

**Table 1.** Safety and tolerability (intention-to-treat, n=20).

| **Electrode-site skin grading (worst grade per session, graded by study staff)** |  |  |  |  |
| --- | --- | --- | --- | --- |
| **Stage** | **No reaction** | **Mild** | **Moderate** | **Severe** |
| Baseline / safety run-in (43 gradings) | 2 | 31 | 10 | 0 |
| Treatment week (157 gradings) | 12 | 125 | 20 | 0 |
| Worst grade per participant (n = 20) | 0 | 12 | 8 | 0 |
| **tDCS adverse-event questionnaire (203 post-session administrations)** |  |  |  |  |
| **Symptom** | **Participants reporting, n of 20** | **Sessions, any ≥ mild, %** | **Sessions, ≥ moderate, %** | **Attributed to device, n of 20** |
| Fatigue / sleepiness | 16 | 49.3 | 20.2 | 3 |
| Burning sensation at/near electrode | 15 | 45.8 | 12.8 | 11 |
| Tingling at/near electrode | 15 | 43.3 | 10.3 | 12 |
| Difficulty concentrating | 16 | 40.4 | 15.8 | 4 |
| Headache | 12 | 34.5 | 8.9 | 9 |
| Itching at/near electrode | 9 | 30.5 | 5.4 | 8 |
| Anxiety / irritability | 11 | 28.6 | 8.4 | 1 |
| Nausea | 10 | 17.2 | 1.0 | 6 |
| Metallic taste | 6 | 17.2 | 3.4 | 6 |
| Mood worsening / dysphoria | 9 | 16.7 | 4.4 | 2 |
| Scalp pain / discomfort | 7 | 15.8 | 3.0 | 6 |
| Dizziness / lightheadedness | 9 | 13.3 | 2.0 | 6 |
| Neck pain | 8 | 11.8 | 0.5 | 2 |
| Visual changes (e.g., phosphenes) | 3 | 4.9 | 0.5 | 1 |

Skin: 200 electrode-site gradings from 203 delivered sessions; no skin breakdown occurred at
any assessment. Adverse-event questionnaire: participants reporting = endorsed the symptom at
any severity in at least one session; attribution to the device was recorded per participant rather
than per session. Symptoms are ordered by per-session endorsement.

### Accelerated HC-tDCS was tolerated on the tDCS Adverse Event Questionnaire

On the tDCS adverse-event questionnaire (203 post-session administrations), the most frequently endorsed symptoms were fatigue or sleepiness (49.3% of sessions; 16 of 20 participants), burning (45.8%; 15 of 20) and tingling (43.3%; 15 of 20) at the electrode, difficulty concentrating (40.4%; 16 of 20) and headache (34.5%; 12 of 20) (Table 1); mean severity across all symptoms and sessions was 1.34 on the 1–5 scale (SD 0.28; CI 1.21 to 1.47), between absent and mild. Symptoms attributed to the device were mainly local sensations: tingling (12 of 20), burning (11), headache (9) and itching (8). No participant moved from absent to present on PHQ-9 item 9 (suicidal ideation) through day 5. Three events were entered in the adverse-event log (two moderate, one mild), none serious (Supplementary Methods 2). The first, judged probably related to stimulation, was twitching around the right eye followed by rhythmic movements of the right arm during the ramp to 6 mA; the participant remained alert and conversant, and the movements stopped as soon as the amplitude was reduced to 4.5 mA and did not recur. The second, judged probably not related, was worsening depression with a passive suicidal remark four days after treatment in a non-responder, managed with safety planning. The third, judged related, was hyperpigmentation at the anode site that persisted unchanged at one month.

Eighteen participants completed an end-of-treatment feedback questionnaire: 14 (78%) would undergo the treatment again and 13 (72%) felt it had helped their symptoms; one participant answered no to both.

### Depressive symptoms improved following accelerated HC-tDCS

Self-rated PHQ-9 scores decreased from 15.8 at baseline to 7.2 at end of treatment (day 5), a mean reduction of 8.6 points (CI 5.5 to 11.6; t(19)=5.84, p<0.001; Hedges g=1.62), or 54.3% (Fig. 2A). Ten of 20 participants (50%; CI 27% to 73%) responded and 9 (45%; CI 23% to 68%) remitted. Response did not differ by self-reported treatment resistance: 8 of the 15 participants reporting two or more failed adequate antidepressant trials responded (53%), compared with 2 of the 5 whose treatment history was unknown (p=1.0, Fisher exact test). In the per-protocol population (n=17), 10 responded and 9 remitted (Supplementary Fig. 2). Improvement accrued progressively across the treatment week (Fig. 2B).

**Figure 2.**
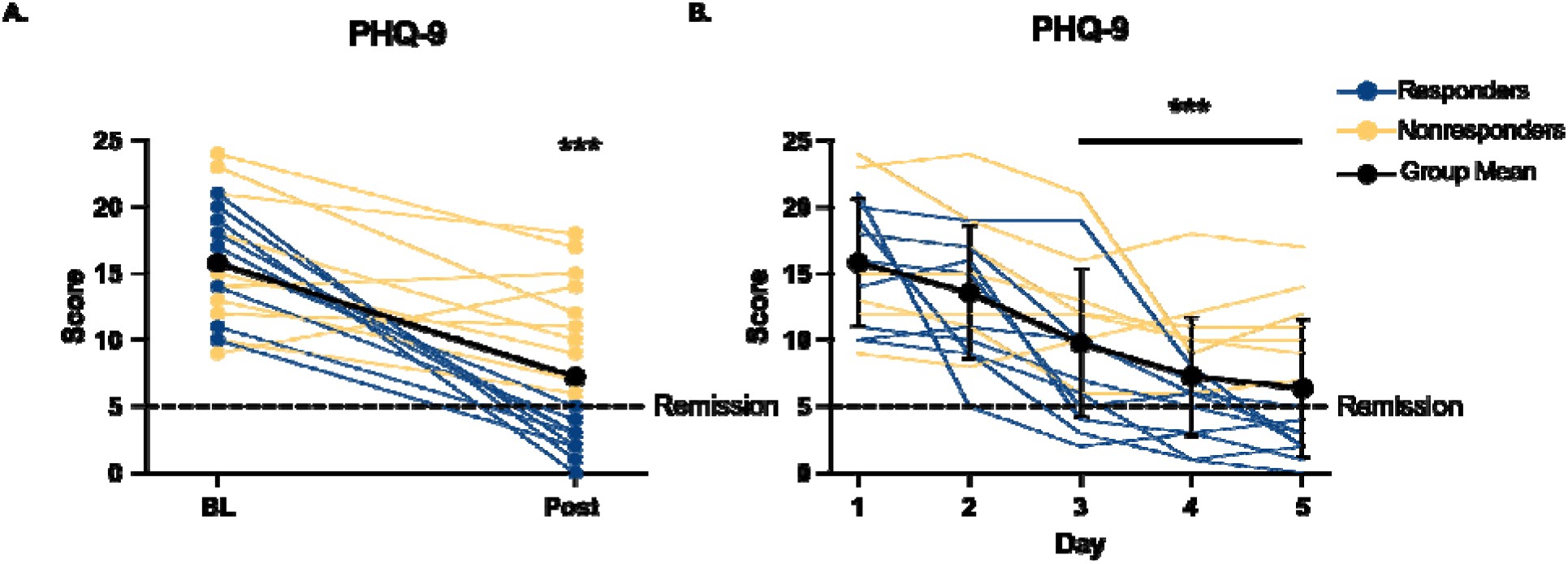
Depressive symptoms decreased progressively over the five-day treatment course. (A) PHQ-9 at baseline (BL) and end of treatment (Post), intention-to-treat (n = 20). Blue and yellow lines are responders (50% or greater reduction) and non-responders; bold line, group mean; dashed line, remission threshold (PHQ-9 below 5). (B) Daily PHQ-9 in the 16 participants with complete daily ratings. A repeated-measures one-way ANOVA indicated a significant effect of day, and post hoc analysis found a significant reduction in scores from day 1 by day 3, maintained at days 4 and 5 (both adjusted p<0.001). Bold line, group mean with SD; thin lines, individual participants colored by response at day 5. A per-protocol version (n=17) is provided as Supplementary Fig. 2.

Pre-treatment expectancy was not significantly associated with symptom change (r=−0.40, p=0.079; Supplementary Fig. 3). In the 7 participants with clinician-rated assessment, MADRS decreased from 28.1 to 17.6 (p=0.022) and HRSD-24 from 27.0 to 20.0 (p=0.058), and both scales tracked self-report closely (Supplementary Fig. 4).

All four secondary instruments improved (Fig. 3): anhedonia on the SHAPS (33.3 to 26.1; p<0.001; g=1.23), mindfulness on the CAMS-R (26.7 to 32.3; p=0.002; g=0.76), self-reported cognitive function on the PROMIS Cognitive Function T-score (33.6 to 44.1; p<0.001; g=1.23), and Trail Making Test parts A (27.7 to 22.5 seconds; p<0.001; g=0.57; n=19) and B (66.3 to 49.6 seconds; p=0.002; g=0.61; n=18). Wilcoxon signed-rank tests agreed for every measure; per-protocol results are given in Supplementary Results.

**Figure 3.**
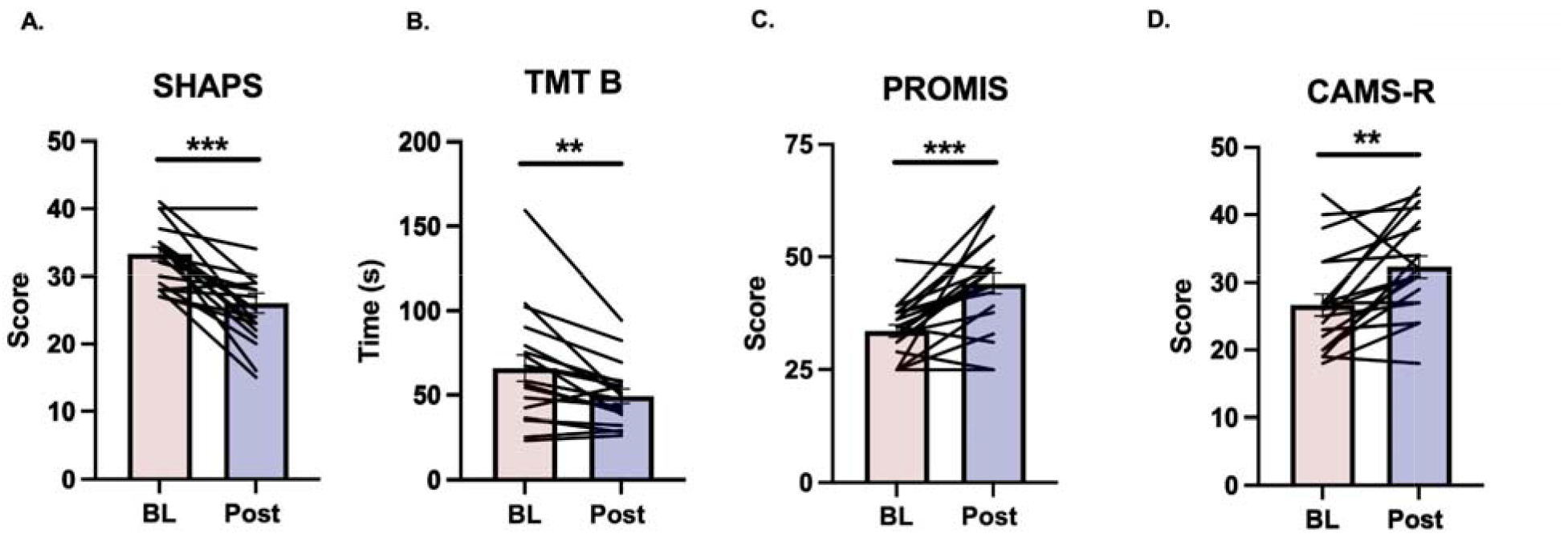
Secondary measures from baseline (BL) to end of treatment (Post), intention-to-treat. Bars are group means with standard error and thin lines are individual participants. (A) SHAPS, anhedonia on the 14–56 scale (lower = better; n = 20). (B) Trail Making Test part B, seconds (lower = better; n = 18). (C) PROMIS Cognitive Function 4a T-score (higher = better; n = 20). (D) CAMS-R, mindfulness (higher = better; n = 20). Paired t tests, **p<0.01, ***p<0.001. Effect sizes are given in the text.

### Improvement was maintained to one month

PHQ-9 improvement was maintained through one month (Fig. 4). A mixed-effects model showed a significant effect of time (F(2.13, 38.40)=24.75, p<0.001), with scores significantly below baseline at end of treatment, one week and one month (Holm–Šídák adjusted p<0.001). Mean PHQ-9 was 7.2, 7.9 and 8.1 at these timepoints (n=20, 20 and 17), and 13 of 20 (65%) met the response criterion at one week. SHAPS, CAMS-R and PROMIS Cognitive Function also remained significantly improved at one week and one month (adjusted p≤0.003; Supplementary Results). Participants were free to change treatments during follow-up.

**Figure 4.**
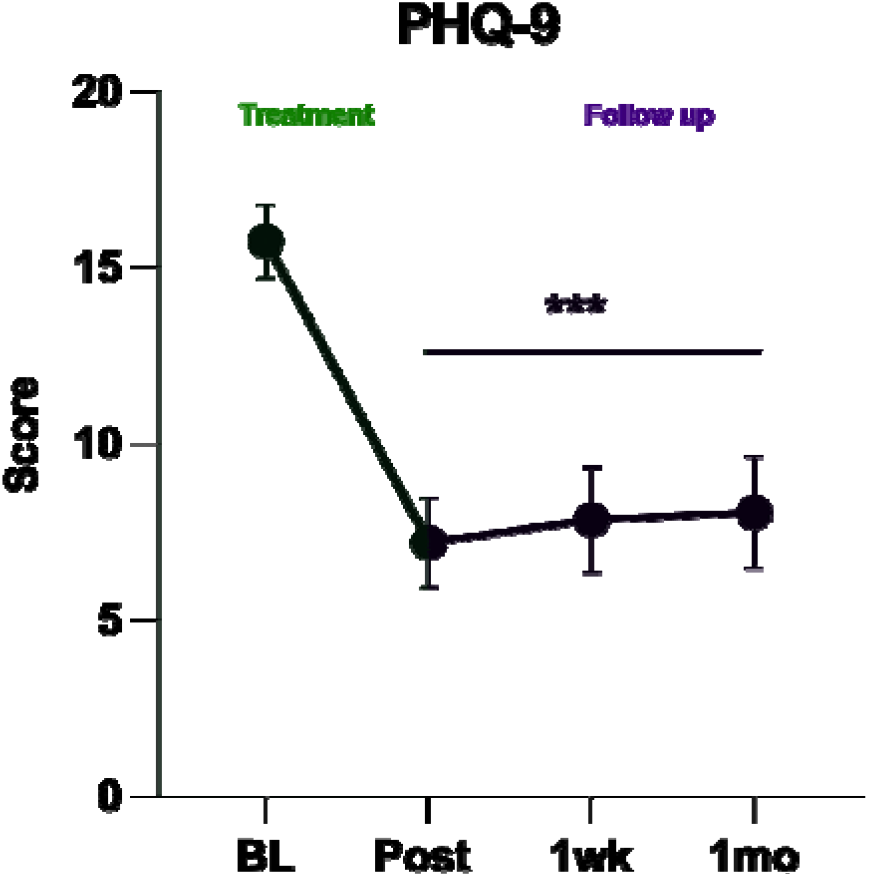
PHQ-9 from baseline through one month of follow-up (intention-to-treat). Points are observed means with standard error (n = 20, 20, 20 and 17). A repeated-measures mixed-effects model with the Greenhouse–Geisser correction showed a significant main effect of time (F(2.13, 38.40)=24.75, p<0.001). Holm–Šídák post hoc comparisons against baseline were significant at end of treatment, one week and one month. ***Adjusted p<0.001 versus baseline.

## Discussion

Accelerated HC-tDCS at 6 mA, three times the amplitude of the FDA-approved at-home protocol, delivered twice daily for five days, was feasible and well tolerated. All 20 participants completed the course, and 78% reported that they would do the treatment again. Amplitude was reduced for intolerance in one participant and limited by an electrode fault in two. Sensations during stimulation were mostly mild and local, chiefly tingling and burning at the electrode. One participant developed twitching around the right eye followed by rhythmic movements of the right arm, limiting him to 4.5 mA.

Skin effects were the most frequent adverse finding: no severe reaction or skin breakdown occurred across 203 combined treatments, though 8 of 20 participants had moderate erythema at least once. The 2 participants with stimulation-related discoloration at one month were the only Black participants enrolled, and the change presented as hyperpigmentation, which can persist for months after inflammation in darker skin^33^ and which erythema-only grading does not capture. Prior accelerated 2 mA tDCS produced irritant contact dermatitis in 62% of participants over two weeks^34^, beyond the duration of our course. Future trials should record Fitzpatrick skin type, document pigment change, include dermatological assessment, and inform participants with darker skin of this risk.

With respect to secondary outcomes in this study, depressive symptoms decreased markedly during the treatment week, with most of the reduction by day 3, and improvement did not differ by reported treatment resistance. Randomized tDCS trials with conventional dose regimes reach peak effects after about six weeks ^25^. Response and remission here, measured by self-report at day 5, were comparable to open-label standard rTMS, which takes 7-8 weeks and over 30 sessions to reach its endpoints ^4,5^. In this study, 50% of participants responded and 45% remitted after ten sessions over five days, and the effects were maintained at the 1-month follow-up, though these are descriptive comparisons, not tests of relative efficacy.

Two tDCS sessions spaced 20 minutes apart interact to prolong the durability of LTP-like plasticity ^27^ potentially allowing effects lasting >24 hour to interact with subsequent treatment days. This cumulative plasticity hypothesis also underlies accelerated TMS protocols, including SAINT ^28,29^ and accelerated 2 mA tDCS ^24^. The combination of optimized electrode placement and increased current amplitude produces a 4.5-fold increase in brain electric field magnitude, achieving >1 V/m across superficial and deep brain regions implicated in mood regulation ^12^. The combination of increased dose intensity and accelerated treatment may produce nonlinear accumulation of therapeutic benefit. Interestingly, within this cohort, the three participants who did not receive the intended 6 mA, whether for tolerability or electrode malfunction, responded worse than the per-protocol group who received the full 6 mA throughout the entire protocol. While exploratory, this is consistent with the clinical value of the full dose regime.

All other secondary measures showed significant improvement by the end of the five-day treatment course, including anhedonia (SHAPS), subjective cognition (PROMIS), executive function (Trail Making Test), and trait mindfulness (CAMS-R). Improvements in anhedonia are particularly relevant given its association with suicidality and poorer antidepressant response ^35–39^, while improvements in both subjective and objective cognitive measures are consistent with previous reports of cognitive benefits from tDCS ^21,26,40–42^. Notably, trait mindfulness also increased despite the absence of explicit mindfulness training, which may be clinically important as mindfulness has been consistently implicated in the treatment of depression, pain conditions, smoking, and addictive disorders ^43–46^. Together, these findings suggest improvements across multiple clinically relevant domains beyond depressive symptom scores, although their close association with overall depression improvement, as well as potential practice effects on cognitive testing, limits conclusions about independent treatment effects.

This study’s notable strengths include being, to our knowledge, the first to combine 6 mA tDCS with an accelerated schedule and having a Safety Review Committee. Retention and adherence were 100%. Safety was measured at every session, and skin was followed for one month, with photographs taken at each timepoint. The study also has limitations. It was open-label without a sham arm, so expectancy, the substantial staff contact of twice-daily in-laboratory sessions, regression to the mean and the natural course of the episode cannot be separated from any effect of stimulation. The primary symptom outcome was self-reported, with clinician ratings in only 7 participants. The sample was small, drawn from a specialty referral stream and predominantly White (85%), and treatment resistance was self-reported. Trail Making gains may reflect practice effects, medications were not monitored after entry, and an electrode fault prevented 6 mA in two participants.

## Conclusion

In the first clinical study of accelerated HC-tDCS, delivering 6 mA, three times the amplitude of the FDA-approved protocol, twice daily for five days was feasible and tolerated, with no serious adverse events and no discontinuations. Depressive symptoms decreased substantially within the treatment week, and the improvement was maintained at one month. Electrode-site skin changes were the principal adverse finding, and pigment change in darker skin should be monitored prospectively. A randomized sham-controlled trial is needed to determine whether these improvements reflect a specific effect of stimulation. If confirmed, accelerated HC-tDCS may prove to be a scalable, rapid-acting brain stimulation intervention, effective even in treatment resistant populations.

## Data availability

The data supporting the findings of this study contain potentially identifying clinical information from a small cohort and are therefore not publicly available. De-identified participant-level data for the reported outcomes, together with the data dictionary, are available from the corresponding author on reasonable request from qualified investigators, subject to a data use agreement and approval by the Medical University of South Carolina Institutional Review Board. Requests will be answered within eight weeks. Aggregate results are reported in full in the manuscript and its supplementary information. The trial is registered at ClinicalTrials.gov under NCT07226011, where the protocol and results will be posted.

## Author contributions

C.O. conceived and designed the study, prepared the regulatory documentation, oversaw study conduct, delivered stimulation sessions, managed project administration, and drafted the manuscript. E.C. performed screening, delivered the majority of stimulation sessions, and conducted follow-up assessments. D.S. contributed to study conception, delivered stimulation sessions, and revised the manuscript. S.C. produced the statistical analyses, advised on statistical methodology and revised the manuscript. M.B. contributed to study conception, provided the investigational device and electrodes through his laboratory and contributed to manuscript drafting/editing. K.D., M.F. and M.B. co-invented the high-capacity electrodes; K.D. and M.B. coordinated device design and trained the study team in device operation. M.B.K. assisted with E-Field processing. J.-P.M. advised on protocol development, contributed to drafting the study protocol and revised the manuscript. C.L. and D.F. assisted with study administration and data collection. J.C. served as a neurological consultant and helped with manuscript editing. M.S.G. provided technical oversight as principal investigator, contributed to study conception, manuscript drafting, and supervised the study.

## Acknowledgements

We thank André R. Brunoni, MD, PhD, and Jeffrey J. Borckardt, PhD, for serving on the Safety Review Committee, and Paul J. Nietert, PhD for statistical consultation. We are grateful to the participants who volunteered for this study.

