## Supplementary Information for "Accelerated high-capacity 6 mA transcranial direct current stimulation for major depression"

### Supplementary Figures and Table

Supplementary Fig 1. removed in accordance with medRxiv policy.

**Supplementary Fig. 1. Electrode montage placement.** Self-adhesive high-capacity hydrogel electrodes in the bifrontal montage, anode over the left and cathode over the right frontal region, each positioned as laterally as possible following the contour of the eyebrow. (A) Frontal view. (B) Oblique view of the left (anode) site. The individual shown is a member of the study team, not a study participant, and provided written informed consent for publication of this image.

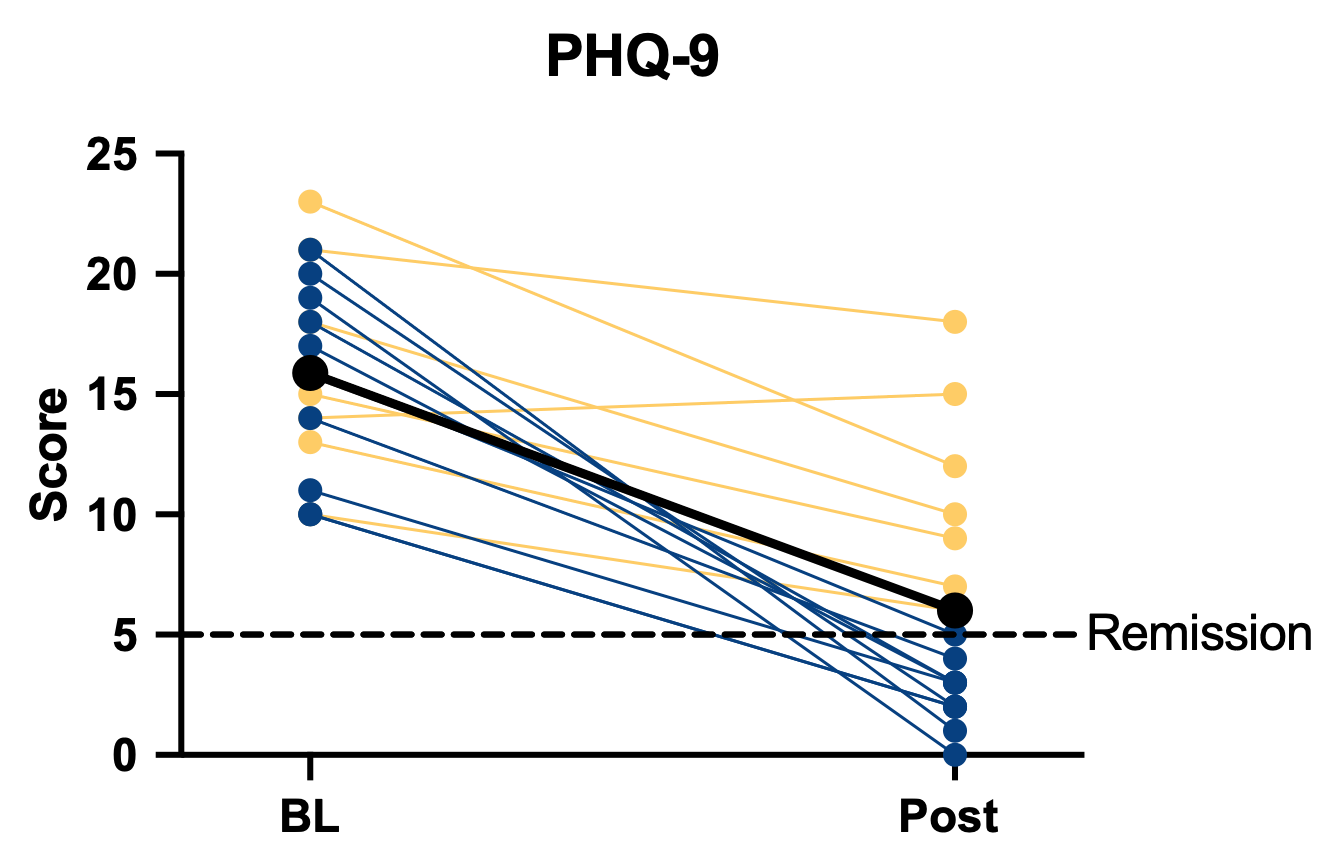

**Supplementary Fig. 2. PHQ-9 from baseline to end of treatment (per-protocol, n=17).** Per-protocol counterpart to Figure 2A. Individual trajectories with responders (50% or greater reduction) and non-responders distinguished; bold line is the group mean. Hedges g=2.05; 10 of 17 participants responded and 9 remitted. The dashed line marks the remission threshold (PHQ-9 below 5).

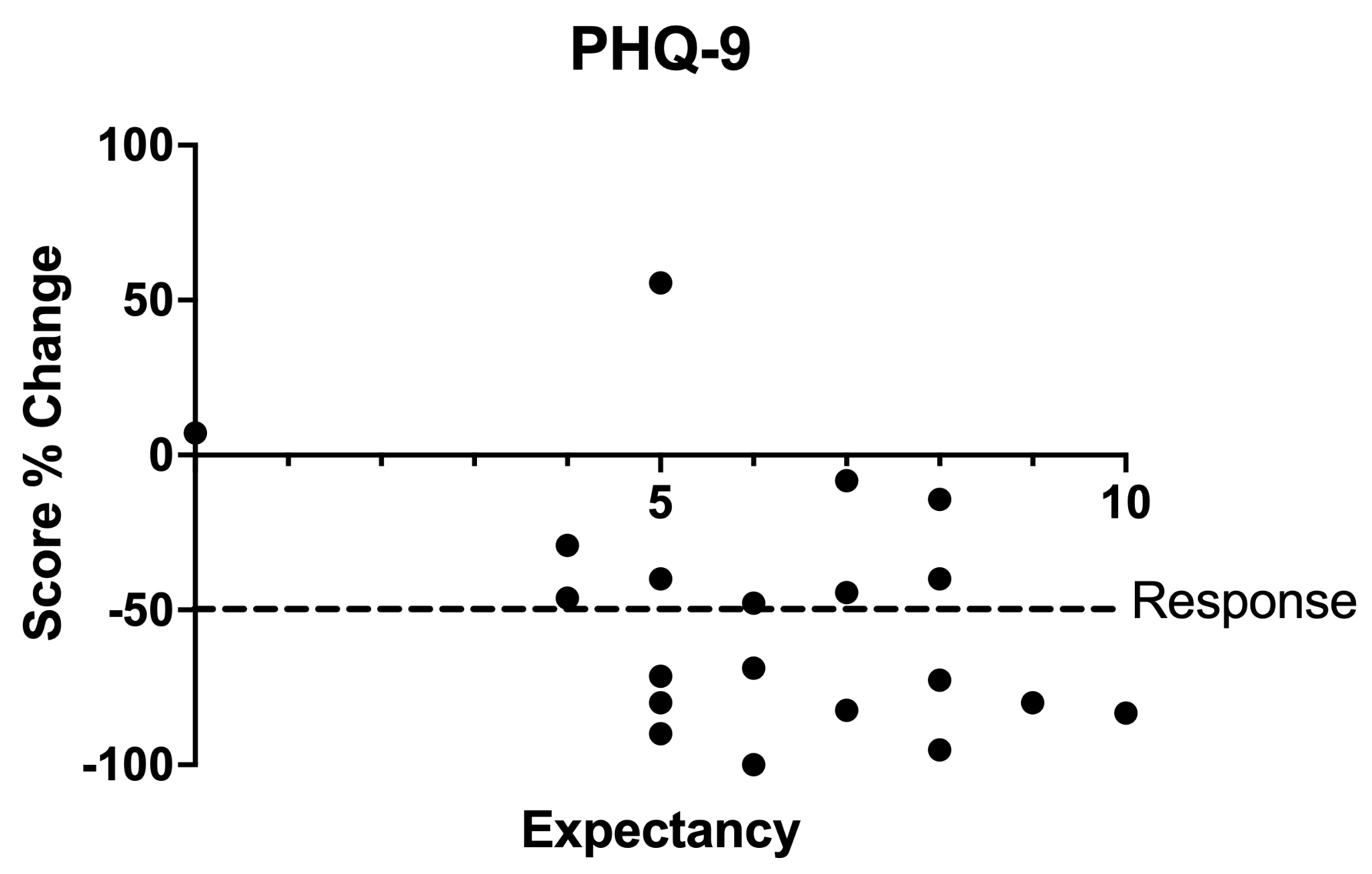

**Supplementary Fig. 3. Pre-stimulation treatment expectancy was not significantly associated with symptom change.** Intention-to-treat, n=20. Expectancy was rated from 0 to 10 before the first stimulation session. Negative values indicate improvement; the dashed line marks the response threshold (50% reduction). Pearson r=−0.40 (CI −0.72 to 0.05, p=0.079);

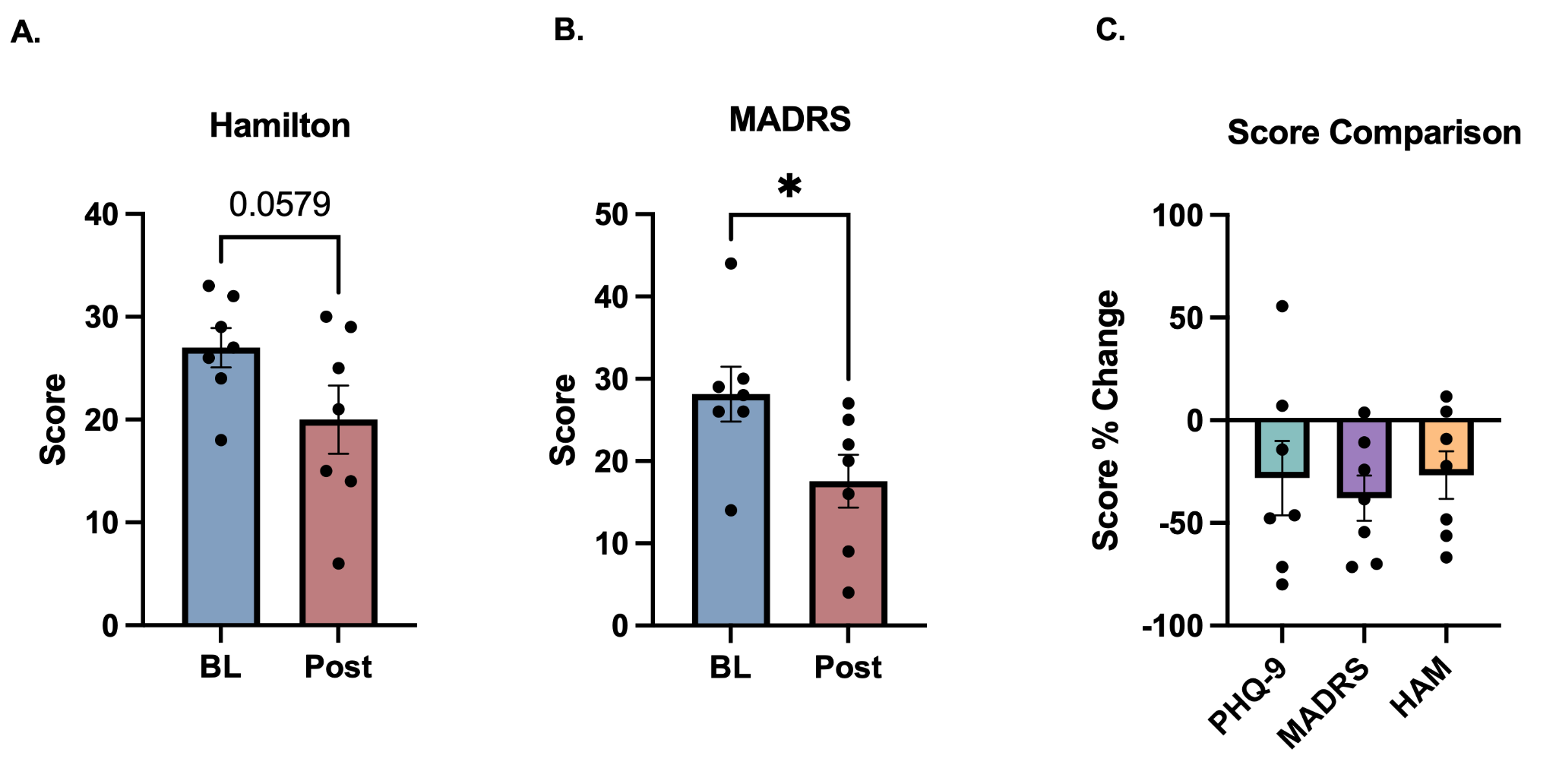

**Supplementary Fig. 4.** Clinician-rated depression scores decreased in parallel with self-report following accelerated HC-tDCS. (A) A paired t-test of HRSD-24 indicated a strong trend in score attenuation from baseline to end of treatment, however it did not reach statistical significance [t (6) = 2.340, p = 0.058]. (B) A paired t-test of MADRS total score indicated a significant reduction over time [t (6) = 3.083, p = 0.022]. (C) A repeated-measures one-way ANOVA of percent change from baseline revealed no significant difference among the clinician-rated scales and the PHQ-9 in the same participants [F (1.19, 7.13) = 1.298, p = 0.302]. Clinician-rated assessment was added by protocol amendment partway through enrollment and was available in 7 of 20 participants.

* p < 0.05.

**Supplementary Table 1. Baseline characteristics (intention-to-treat, n=20).** Percentages are of the 20 participants who received at least one stimulation session. Adequate prior antidepressant trial is defined as at least six weeks at a therapeutic dose, by participant report.

| **Characteristic** | **Value** |
| --- | --- |
| Age, years, mean (SD) | 39.6 (14.9) |
| Age, range | 18 to 69 |
| Sex assigned at birth, female | 11 (55%) |
| Gender identity, woman / man / non-binary | 9 / 9 / 2 |
| Race, White | 17 (85%) |
| Race, Black or African American | 2 (10%) |
| Race, not reported | 1 (5%) |
| Ethnicity, not Hispanic or Latino | 20 (100%) |
| Duration of current episode, more than 12 months | 8 (40%) |
| Duration of current episode, 6 to 12 months | 7 (35%) |
| Duration of current episode, 3 to 6 months | 3 (15%) |
| Duration of current episode, 1 to 3 months | 1 (5%) |
| Duration of current episode, less than 1 month | 1 (5%) |
| Baseline PHQ-9, mean (SD) | 15.8 (4.7) |
| Baseline PHQ-9, range | 9 to 24 |
| Two or more adequate prior antidepressant trials | 15 (75%) |
| Currently taking an antidepressant | 12 (60%) |
| Psychiatric medication change in prior 2 months | 2 (10%) |

### Supplementary Methods

#### Supplementary Methods 1. Detailed methods

**Eligibility.** Adults aged 18–70 years were eligible if they had a current diagnosis of major depressive disorder established by the Mini-International Neuropsychiatric Interview version 7 (MINI v7.0.2), a baseline PHQ-9 score greater than 9, capacity to provide informed consent, and spoke fluent English. Exclusion criteria were bipolar or psychotic disorder; a primary anxiety disorder without concomitant major depression; current significant suicidal ideation or behavior requiring a higher level of care; use of neuromodulation therapy (electroconvulsive therapy, transcranial magnetic stimulation, or vagus nerve stimulation) within the preceding six months; history of seizure, implanted cranial or cardiac metal, or neurosurgery; use of medications that significantly reduce the seizure threshold; catatonia or other inability to complete the consent process; frequent or severe headache; any contraindication to MRI; personal history of head trauma, concussion, or traumatic brain injury; current moderate-to-severe alcohol or substance use disorder; any non-uniformity of the skin at the electrode sites, including eczema, rash, hyperhidrosis, communicable skin disorder, blister, open wound, burn, or laceration, as determined by clinical personnel; and pregnancy. Women of childbearing potential underwent urine pregnancy testing at screening and before stimulation.

**Stimulation device and electrodes.** Stimulation was delivered with a High-Capacity 1×1 transcranial direct current stimulator (HC-1×1; Soterix Medical), a customized version of the Soterix Medical 1×1 platform. Three modifications distinguish it from the standard device: a maximum output extended to 6 mA, an adaptive current ramp lengthened from 60 to 120 seconds, and a maximum compliance voltage of 7.5 V, approximately five-fold below that of the standard platform. The modified device is investigational and is not approved by the FDA for the treatment of any condition.

Current was delivered through self-adhesive high-capacity hydrogel electrodes manufactured in the laboratory of M.B. (City College of New York). Each electrode had a 25 cm² contact surface, matching the area of a conventional 5 × 5 cm sponge pad. Electrodes were placed in a bifrontal montage with the anode over the left frontal region and the cathode over the right frontal region, each positioned as laterally as possible following the contour of the eyebrow (Supplementary Fig. 1). Electrodes were single-use and were replaced for every session.

**Session delivery.** Each stimulation session delivered 6 mA for 20 minutes. Participants received two sessions per day, separated by a 30-minute rest interval, on five consecutive weekdays, for a target of 10 sessions and 200 minutes of stimulation at 6 mA per participant; the three safety run-in participants received one additional Day 1 session during amplitude escalation. The cohort therefore received 203 sessions in total. Delivered current was not logged by the device; any session in which the 6 mA target was not sustained was recorded by study staff in the session record, and these records define the sub-target subgroup reported in Results. All sessions were delivered in the laboratory with a co-investigator present and continuous staff monitoring. Participants remained seated throughout stimulation but were given no concurrent task or instruction: some read, some listened to music, and some sat quietly. No cognitive training, psychotherapy, or other structured activity was paired with stimulation, and concurrent activity was not recorded.

**Skin assessment.** Before stimulation each day, study staff inspected both electrode sites and recorded any finding, its side, its type (discoloration, acne, blemish, dryness or flaking, irritation or rash, cut, open skin, other), its severity, and any action taken, such as repositioning the electrode. After every session, staff graded erythema at the more affected site (none, mild, moderate, severe). The daily pre-stimulation check was introduced after the first safety run-in participant. Non-identifying photographs of the electrode sites were taken before stimulation each day and after each session. At each follow-up visit, participants completed a questionnaire on electrode-site skin changes (discoloration, dryness, itching, redness, flaking, tenderness, pain), rated change since the last visit, and uploaded forehead photographs. Photographs from participants were reviewed by one unblinded investigator (C.O.), not a dermatologist.

**Clinician-rated assessment, expectancy and MRI.** Clinician-rated assessment was added by protocol amendment partway through enrollment, after the early self-report changes proved larger than anticipated; it yields HRSD-24 and MADRS totals from a single interview, administered at baseline and end of treatment. Coverage is partial, and it was not a prespecified outcome.

Treatment expectancy was rated on a single 0–10 item (0, not expecting any benefit; 10, extremely confident it will help) immediately before each stimulation session. The rating obtained before the first session, before any stimulation, was used as the pre-treatment measure.

As a pre-specified exploratory outcome, all participants were offered a structural T1-weighted MRI for retrospective individualized electric-field modeling. Modelled field values and their relationship to outcome are reserved for a separate report.

**Assessment schedule and scoring conventions.** Baseline was taken as the assessment obtained on the first treatment day, immediately before the first stimulation session, rather than the screening visit. The PHQ-9 was administered at three recall periods: the standard instrument at screening, baseline, and the post-intervention visit and every follow-up beyond one week; an intersession instrument during the treatment week; and a dedicated instrument at the one-week follow-up. The three instruments differed only in the instructions as to the window of time that asked the patient to assess their symptom severity (e.g., the standard PHQ-9 instructs patients to report symptoms over the past two weeks, the intersession directs them to the past 24 hours, and the one-week follow-up refers to the prior week). Baseline came from the standard instrument for all 20 participants. The Day 5 value was taken from the final intersession administration within the treatment week for 18 participants and from the standard instrument for the two earliest enrolled. Where a PHQ-9 total was missing but all nine items were present, the total was reconstructed as the item sum and flagged as reconstructed.

CAMS-R items 2, 6, and 7 were reverse-scored (5 − raw) before summing, as the instrument requires. SHAPS was scored on the 14–56 continuous Likert scale, with higher scores indicating greater anhedonia, such that improvement appears as a decrease. PROMIS Cognitive Function 4a raw scores were converted to T-scores using the official conversion table, referencing each score to a population mean of 50 and standard deviation of 10.

**Analysis populations.** Outcomes are reported in three nested populations: Intention-to-treat (ITT): all participants who received at least one stimulation session. Completer: participants whose delivered sessions spanned at least five distinct treatment days (10 sessions). Per-protocol: completers who sustained the full 6 mA target amplitude throughout. ITT is the primary population for all efficacy outcomes. The per-protocol population is reported alongside it to address the narrower question of what occurs when the intended dose is delivered. Safety and tolerability are reported exclusively on the ITT population. No safety analysis is split by population, since excluding an exposed participant from a safety denominator would understate risk. Participants who consented but never began stimulation are screen failures, sit outside all three populations, and are not imputed.

**Statistical analysis.** The study was designed to estimate safety, tolerability and feasibility and to generate effect-size estimates for a subsequent randomized trial. It was not powered to test efficacy and no formal power calculation for a treatment effect was performed. A sample of 20 was chosen pragmatically to estimate tolerability and feasibility at 6 mA, not to test efficacy. All efficacy analyses are therefore estimation-focused and are interpreted as within-participant change rather than as treatment effects: in the absence of a sham arm, no analysis reported here can separate the effect of stimulation from expectancy, regression to the mean, or the natural course of the episode. No statistical analysis plan was prespecified. The protocol prespecified the primary safety and tolerability outcomes, the secondary instruments and the assessment schedule; the analytic conventions reported below, including the response and remission thresholds, were adopted after data collection. All symptom outcomes are exploratory, as the protocol states.

For each outcome measure, several statistical methods were used to characterize the response to accelerated tDCS in this population. First, the mean change from baseline to end of treatment (day 5) was calculated for each measure and compared using two-sided paired t-tests, with Wilcoxon signed-rank tests performed alongside as a distribution-free sensitivity analysis. These values were used to calculate Hedges' g effect sizes on the pooled baseline and end-of-treatment standard deviation. Next, a repeated-measures one-way ANOVA (or a mixed-effects model where follow-up data were missing) was performed across baseline, end of treatment, one week, and one month in order to assess whether the change from baseline was durable. Sphericity was assessed, and Greenhouse–Geisser corrections were applied as needed. Daily PHQ-9 across the treatment week was analyzed with the same repeated-measures one-way ANOVA, with each day compared against day 1. Finally, response was defined as a reduction of at least 50% in PHQ-9 and remission as a PHQ-9 score below 5; these and the safety proportions are reported with exact (Clopper–Pearson) binomial confidence intervals. Adverse events and skin grades were tabulated descriptively. Pre-stimulation findings and follow-up skin reports were counted per participant. The association between pre-treatment expectancy and percent change in PHQ-9 was assessed with Pearson and Spearman correlations. In the subset with clinician-rated assessment, HRSD-24 and MADRS were compared from baseline to end of treatment using paired t-tests, percent change on the two clinician scales and the PHQ-9 was compared by repeated-measures one-way ANOVA, and their agreement with the PHQ-9 was summarized by a within-participant correlation computed on participant-centered scores. All post hoc analysis was performed using the Holm–Šídák multiple comparisons test, all t-tests were two-sided with α = 0.05, and all confidence intervals are 95%. Analyses were performed using GraphPad Prism (v11).

#### Supplementary Methods 2. Adverse-event narratives

The first, judged probably related to stimulation, occurred during the ramp to 6 mA: twitching around the right eye, followed by rhythmic movements of the right arm. No movement of the leg was observed, although the participant reported a sensation in the right leg. The participant remained alert and conversant throughout. The movements stopped as soon as the amplitude was reduced to 4.5 mA, with no subsequent confusion, fatigue or other symptoms, and they did not recur at 4.5 mA for the remainder of that participant's course.

The second, judged probably not related, occurred four days after the final session in a participant who had not responded to treatment (PHQ-9 14 at baseline, 15 at day 5): a family member reported worsening depressive symptoms and a passive suicidal remark. The participant was safety-planned and referred to outpatient care and required no emergency evaluation or hospitalization; the history obtained for this patient included a similar deterioration following esketamine and following medication changes in the past.

The third, judged related to stimulation, was hyperpigmentation at the anode site that was present at one week and persisted, unchanged, at one month; it caused no symptoms and required no treatment, and follow-up is ongoing.

### Supplementary Results

**Enrollment and participants.** Between 15 December 2025 and 28 August 2026, 29 individuals were assessed for eligibility and 25 provided written informed consent, consent having been obtained before final eligibility determination in some cases; 9 were not enrolled (8 ineligible and 1 who withdrew after consent) and 20 received at least one stimulation session (Fig. 1). These 20 participants constitute the intention-to-treat population. After the three safety run-in participants completed treatment, the Safety Review Committee met, reviewed their adverse-event, skin and tolerability data, and determined that enrollment could proceed at 6 mA. All 20 completed the full five-day course, so the completer population is identical to the intention-to-treat population. Three participants did not sustain the 6 mA target for the full course: one because of intolerance (amplitude reduced to 4.5 mA; see adverse events below) and two because an electrode fault prevented the device from reaching 6 mA. All three were excluded from the per-protocol population (n=17). Mean age was 39.6 years (range 18 to 69) and 11 of 20 were female at birth. Fifteen of the 20 participants (75%) reported two or more adequate prior antidepressant trials. Twelve participants (60%) were taking an antidepressant at entry; two (10%) reported a change in psychiatric medication dose, or a new psychiatric medication, in the two months before enrollment. Baseline characteristics are summarized in Supplementary Table 1.

**Skin outcomes.** Safety and tolerability were the primary outcomes. No participant experienced a device-related serious adverse event (0 of 20; CI 0% to 17%), and all 20 completed every scheduled session without discontinuation for an adverse effect (100%; CI 83% to 100%). Electrode sites were formally graded at 200 of the 203 delivered sessions (98%, 3 were missing a recorded grade in REDCap), and no severe skin reactions or skin breakdown were observed in any session (Table 1). The highest grade recorded for each participant was mild in 12 and moderate in 8. Of the 157 treatment-week gradings, 12 showed no reaction, 125 were mild and 20 were moderate; the remaining 43 were recorded at the baseline visit and during the safety run-in (Table 1). Before stimulation on days 2 to 5, staff recorded a finding at the electrode site in 10 of 19 participants, most often dryness or flaking (7), blemishes (4), discoloration (4) and irritation or rash (3), on the left in 8 and the right in 7. All were mild except one moderate finding. Electrodes were repositioned on at least one day in 5 participants. Four participants had findings on day 1 before any stimulation. At one week, 7 of 20 participants reported a current skin change: discoloration in 5 (mild in 4, moderate in 1), flaking in 4, redness in 3, dryness in 2 and itching in 1. None reported tenderness or pain. A further 5 reported discoloration that had resolved 1 to 5 days after the final session. At one month, 3 of 18 reported residual discoloration (mild in 2, moderate in 1) and no other symptoms. On photographic review, 2 of these 3 cases were judged related to stimulation and 1 judged not related, as the spots were visible on earlier study photographs. The 1 case of discoloration self-rated as moderate and still present at 1 month was reported as an adverse event.

**Adverse-event questionnaire and adverse-event log.** On the tDCS adverse-event questionnaire, administered after every stimulation session rather than at the endpoint of each day (203 administrations per symptom, one per delivered session), the most frequently endorsed symptoms at any severity were fatigue or sleepiness (49.3% of sessions; 16 of 20 participants), burning at or near the electrode (45.8%; 15 of 20), tingling at or near the electrode (43.3%; 15 of 20), difficulty concentrating (40.4%; 16 of 20) and headache (34.5%; 12 of 20) (Table 1). Symptoms the participant attributed to the device were dominated by the expected local sensations: tingling (12 of 20 participants), burning (11 of 20), headache (9 of 20) and itching (8 of 20). No participant moved from absent to present on PHQ-9 item 9 suicidal ideation through day 5. Mean adverse-event questionnaire severity was 1.34 on the 1–5 Likert scale (SD 0.28; CI 1.21 to 1.47), between absent and mild. No severe or extreme symptom was accompanied by an adverse-event log entry, and none required intervention. Three events were entered in the formal adverse-event log, two graded moderate, one mild, and none meeting the protocol definition of a serious adverse event. The first, judged probably related to stimulation, was twitching around the right eye followed by rhythmic movements of the right arm during the ramp to 6 mA. The participant remained alert and conversant, the movements stopped as soon as the amplitude was reduced to 4.5 mA, and they did not recur at that amplitude (Supplementary Methods 2). The second, judged probably not related, occurred four days after the final session in a participant who had not responded to treatment: a family member reported worsening depressive symptoms and a passive suicidal remark. The participant was safety-planned and referred to outpatient care, and required no emergency evaluation or hospitalization (Supplementary Methods 2). The third, judged related to stimulation, was hyperpigmentation at the anode site that was present at one week and persisted, unchanged, at one month; it caused no symptoms beyond appearance and was not given treatment; follow-up is ongoing.

**PHQ-9.** Self-rated PHQ-9 depression scores decreased from 15.8 at baseline to 7.2 at end of treatment (day 5) in the ITT population, a mean reduction of 8.6 points (CI 5.5 to 11.6; t(19)=5.84, p<0.001; Hedges g=1.62). The group mean decreased by 54.3% (Fig. 2A). Ten of 20 participants (50%, CI 27% to 73%) met response criteria at 50% or greater reduction, and 9 of 20 (45%, CI 23% to 68%) met remission criteria at PHQ-9 below 5. Response did not differ by self-reported treatment resistance: 8 of the 15 participants reporting two or more failed adequate antidepressant trials responded (53%) and 7 remitted, compared with 2 of the 5 whose treatment history was unknown (p = 1.0, Fisher exact test). In the per-protocol population (n=17) the reduction was 9.9 points (CI 6.9 to 12.9; t(16)=7.00, p<0.001; g=2.05), with 10 of 17 responders (58.8%) and 9 of 17 remitters (52.9%) (Supplementary Fig. 2). Within the treatment week, improvement accrued progressively rather than at a single point (Fig. 2B).

**Expectancy and clinician-rated assessment.** Pre-stimulation treatment expectancy, recorded before any stimulation was delivered, was not significantly associated with symptom change (r=-0.40, CI -0.72 to 0.05, p=0.079, n=20); the analysis is underpowered at this sample size and is reported in full in Supplementary Fig. 3. Clinician-rated assessment, added partway through enrollment, was available for only 7 of the 20 participants. In those seven, HRSD-24 decreased from 27.0 to 20.0 (t(6)=2.34, p=0.058) and MADRS from 28.1 to 17.6 (t(6)=3.08, p=0.022), against a PHQ-9 fall from 14.9 to 10.3 in the same participants (t(6)=2.05, p=0.087), reductions of 26%, 38% and 31% respectively (Supplementary Fig. 4). Within participants, both clinician scales tracked self-report closely across all 23 administrations (HRSD-24 r=0.835; MADRS r=0.861; both df=15, p<0.001).

**Secondary measures.** All four secondary instruments moved in the direction of improvement (Fig. 3) (ITT values, n=20 unless noted). Anhedonia fell 7.2 points on the SHAPS (33.3 to 26.1; t(19)=5.35, p<0.001, g=1.23; per-protocol 8.4 points, t(16)=6.00, p<0.001, g=1.53); mindfulness rose 5.6 points on the CAMS-R (26.7 to 32.3; t(19)=3.57, p=0.002, g=0.76; per-protocol 6.4 points, t(16)=3.55, p=0.003, g=0.82); self-reported cognitive function rose 10.5 T-score points on the PROMIS Cognitive Function short form, just over one population standard deviation (33.6 to 44.1; t(19)=4.82, p<0.001, g=1.23; per-protocol 12.3 points, t(16)=5.58, p<0.001, g=1.50). Processing speed and set-shifting improved on the Trail Making Test, with part A falling from 27.7 to 22.5 seconds (t(18)=3.95, p<0.001, g=0.57, n=19; per-protocol 28.0 to 23.1, t(15)=3.14, p=0.007, g=0.49, n=16) and part B from 66.3 to 49.6 seconds (t(17)=3.61, p=0.002, g=0.61, n=18; per-protocol 67.9 to 48.9, t(14)=3.69, p=0.002, g=0.63, n=15); the reduced counts reflect missing post-treatment administrations. Wilcoxon signed-rank tests, reported as a distribution-free sensitivity analysis, agreed with the paired t-test for every measure, including the PHQ-9 (all p ≤ 0.002).

**Durability through one month.** Study enrollment concluded on 28 August 2026, on reaching the target of 20 participants; participants were free to start or stop medications and other treatments during follow-up. PHQ-9 was compared across baseline, end of treatment, one week and one month using a repeated-measures mixed-effects model (Fig. 4). A significant main effect of time was observed [F (2.13, 38.40) = 24.75], p<0.0001 (per-protocol [F (2.27, 34.07) = 33.36], p<0.0001). Post hoc analysis indicated a significant reduction from baseline at end of treatment, at one week and at one month following Holm–Šídák correction (largest adjusted p<0.001; per-protocol p<0.001). Observed ITT means were 7.2, 7.9 and 8.1 at end of treatment, one week and one month, against per-protocol means of 6.0, 6.8 and 6.4. Matching by participant accounted for a significant share of the variance [chi-squared (1) = 34.07], p<0.0001. In equivalent models a significant main effect of time was observed for each of the three self-report secondary measures (SHAPS [F (1.72, 29.84) = 13.83], p<0.001; CAMS-R [F (1.55, 27.34) = 10.69], p<0.001; PROMIS Cognitive Function [F (1.72, 30.38) = 15.48], p<0.0001). Post hoc analysis indicated a significant difference from baseline at end of treatment, at one week and at one month for all three measures following Holm–Šídák correction (largest adjusted p=0.003). One-month means were 24.3 on the SHAPS, 33.4 on the CAMS-R and 44.1 on the PROMIS Cognitive Function T-score, against 26.1, 32.3 and 44.1 at end of treatment.

Follow-up is incomplete by design, with later timepoints reached only by earlier enrollees: ITT counts were 20 at baseline and end of treatment, 20 at one week and 17 at one month. Among the 20 participants assessed one week after the course, 13 (65.0%) met the response criterion and 7 (35.0%) the remission criterion.
